# Salzburg Visual Field Trainer (SVFT): A virtual reality device for (the evaluation of) neuropsychological rehabilitation

**DOI:** 10.1101/2021.03.25.21254352

**Authors:** Michael Christian Leitner, Dirk Christoph Gütlin, Stefan Hawelka

**Affiliations:** Centre for Cognitive Neuroscience (CCNS), University of Salzburg, Austria; Department of Psychology, University of Salzburg, Austria; Institute of Cognitive Science, University of Osnabrück, Germany

**Author notes:** Corresponding author: Michael C. Leitner, Ph.D. / University of Salzburg / Centre for Cognitive Neuroscience / Department of Psychology / Hellbrunnerstrasse 34 / 5020 Salzburg / Austria /. The following study was reviewed and approved by the ethics commission of the University of Salzburg (Reference No. 39/2018). The following study is based on the funded project “Advanced Perimetry for the Evaluation of Neuroplasticity in the Visual Cortex” by the Austrian Science Fund (FWF) (Reference No. P31299).

**Keywords:** visual field defect, neuroplasticity, rehabilitation, neuropsychology, virtual reality

## Abstract

**Objective:** So-called “Visual Restitution Therapies” (VRT) claim to ameliorate visual field defects of neurological patients by repeated visual light stimulation, leading to training-related neuroplasticity and resulting in reconnection of lesioned neurons in early cortical areas. Because existing systems are stationary, uncomfortable and unreliable, we developed a training instrument based on virtual reality goggles. The goal of the “Salzburg Visual Field Trainer” (SVFT) is twofold: (1) The device facilitates the clinical evaluation of established neuropsychological rehabilitation approaches, such as VRT. (2) The device enables patients to independently perform VRT based (or other) neuropsychological training methodologies flexibly, comfortably and reliably.

**Methods and Analysis:** The SVFT was developed on the principles of VRT. Individual configuration of the SVFT is based on perimetric data of the respective patient’s visual field. To validate the utmost important procedure in neuropsychological rehabilitation methodologies - that is displaying stimuli precisely in desired locations in the user’s visual field - two steps were conducted in this proof-of-concept study: First, we assessed the individual “blind spots” location and extent of 40 healthy, normal sighted participants. This was done with the help of our recently developed and validated perimetric methodology “Eye Tracking Based Visual Field Analysis” (EFA). Second, depending on the individual characteristics of every participant’s blind spots, we displayed - with the help of the SVFT - 15 stimuli in the respective locations of every participants’ blind spots and 85 stimuli in the surrounding, fully intact visual area. The ratio between visible and non-visible stimuli, which reflects in the documented behavioral response (clicks on a remote control) of the 40 participants, provides insight into the accuracy of the SVFT to display training stimuli in areas desired by the investigator. As the blind spot is a naturally occurring, absolute scotoma in human vision, we utilized this blind area as an objective criterion and a “simulated” visual field defect to evaluate the (technical) methodology of SVFT.

**Results:** Outcomes indicate that the SVFT and its methodology is highly accurate in displaying training stimuli in desired areas of the user’s visual field with an accuracy of 99.0%. Data analysis further shows a sensitivity of .980, specificity of .992, positive predictive value of .955, negative predictive value of .996, hit rate of .990, random hit rate of .742 and RATZ-Index of .976. This translates to 14.7% correct non-reactions, 0.7% false non-reactions, 0.3% false reactions and 84.3% correct reactions to displayed test stimuli during the evaluation study with the SVFT. Reports from participants further indicate that the SVFT is comfortable to wear and intuitive to use.

**Conclusions:** The SVFT can help to investigate the true effects of VRT based methodologies (or other neuropsychological approaches) and the underlying mechanisms of training-related neuroplasticity in early regions of the visual cortex in neurological patients suffering from visual field defects.

## Introduction

Visual field defects are caused by damage to early visual areas in the brain’s occipital cortex and a common consequence of stroke or trauma. Damage in these post chiasmatic areas of the brain usually lead to partial loss of vision - affecting approximately 12% of patients suffering from traumatic brain injury and 35% of patients suffering from stroke (Birnbaum, Hackley & Johnson 2015). The extent of the visual field defect coheres with location and dimension of the cortical lesion. Most common forms are blindness in half (hemianopia) and in a quarter (quadrantanopia) of the visual field (Niedeggen & Jörgens 2005).

So called “Visual Restitution Therapies” (VRT) postulate that damaged areas in the visual cortex can be reactivated by repeated visual stimulation (e.g. Sabel, Kruse, Wolf & Guenther 2013). By presenting bright light-impulses on a computer-screen in the individual transition zone of the intact and defect visual fields, neurons are stimulated and form new connections among residual cortical structures (see Fig. 1).

**Figure 1.**
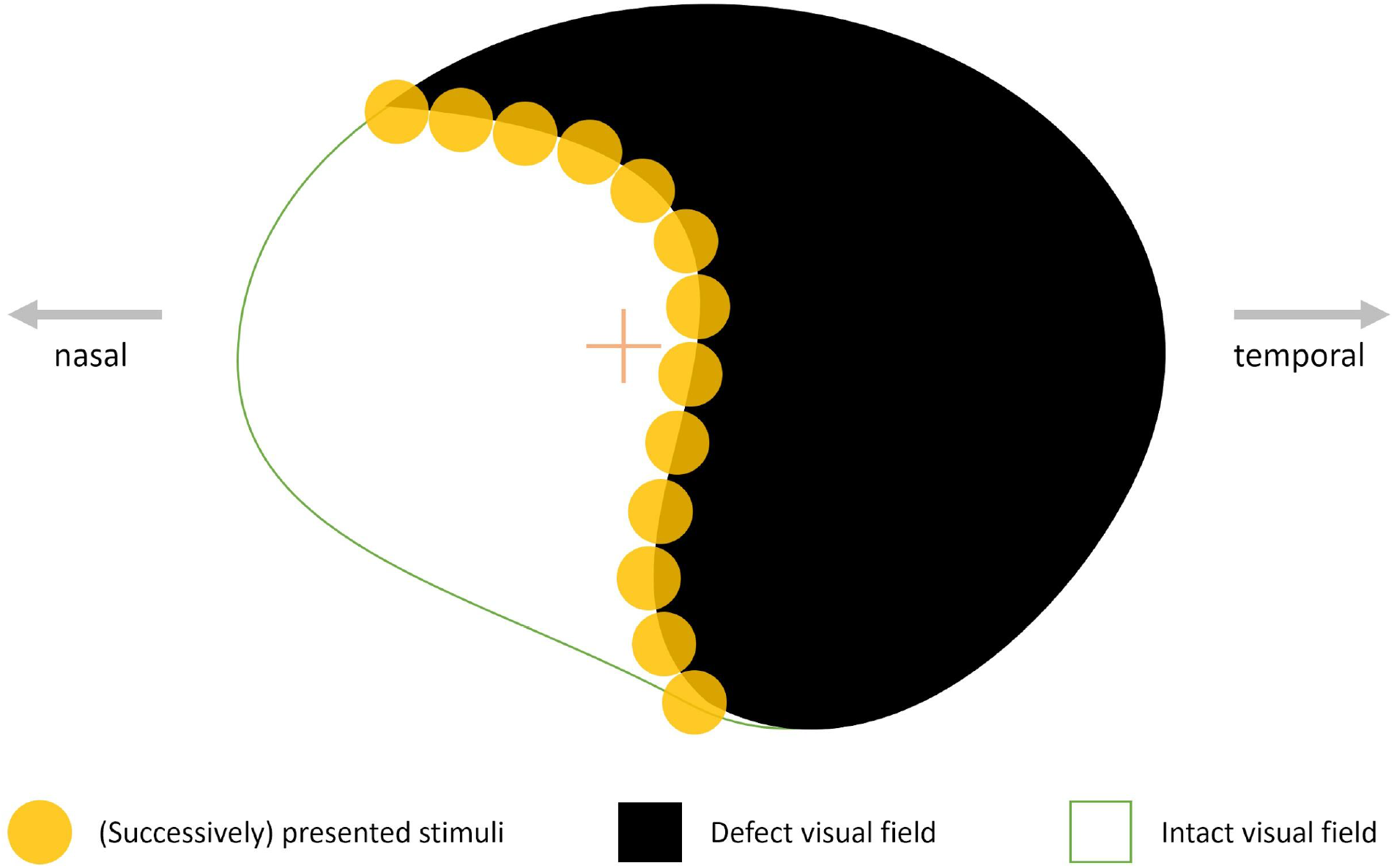
Exemplary illustration of the therapeutic principle of “Visual Restitution Therapies” (VRT). Bright light stimuli are presented randomly across the “border area”, which is between the patient’s defect and intact visual field. Proponents of this therapeutic approach argue that by repeating this process, damaged neurons - due to stroke or trauma - in early visual cortical areas reconnect over time, resulting in a diminished defect and enhanced intact visual field.

Animal studies indicate that damaged neurons in the visual cortex can indeed recover and reconnect by bright light stimulation (Schweigart & Eysel 2002, Huxlin & Pasternak 2004). However, existing evidence for “training-induced neuroplasticity” in the human primary visual cortex is inconsistent. While some studies found a diminution of the blind field after intervention (e.g. Bergsma, Elshout & van den Berg 2017, Marshall et al. 2010; Mueller, Mast & Sabel 2007; Sabel, Kenkel & Kasten 2005), other studies found no significant amelioration and attribute improvement after training to compensatory eye movements during perimetric pre-, post-, and follow-up assessments (e.g. Frolov et al. 2017; Glisson 2006; Horton 2005; Reinhard et al. 2005).

To achieve robust and credible evidence for the effects of interventions based on the principles of VRT and for the issue of training-related neuroplasticity within the human visual cortex, we developed an eye-tracking based perimetric methodology (“Eye Tracking Based Visual Field Analysis” [EFA]) (Leitner et al. 2021). The EFA is based on static automated perimetry and additionally compensates individual eye movements during diagnosis in real time while providing a standard error of measurement of 0.44° of visual angle. This eye-tracking based feature precludes that the diagnosis is unduly influenced by (involuntary) compensation strategies such as (micro) saccades during pre-, post-, and follow-up diagnosis. Study results from patients suffering from cortical lesions (and glaucoma) further indicate that the EFA is applicable for clinical use. Thus, the unprecedented accuracy of the EFA and its insusceptibility to compensation strategies makes the methodology an ideal choice for upcoming investigations regarding results from studies, which found significant amelioration after intervention with VRT based approaches, ascribing post-intervention perimetric changes to the successful stimulation of neuroplasticity in the visual cortex.

In order to realize an evaluation study of VRT based approaches (and other neuropsychological methodologies based on compensatory eye movements in the near future), we sought to develop an ambulant training instrument. The existing concepts and available instruments are coded for Personal Computers and in this way additionally require the purchase of a chin and head rest. This makes therapy inflexible as it is place-bound to a stationary personal computer. Therefore, our newly developed VRT based instrument should be - besides being reliable - portable and easy to use in order to instill high compliance in the trainee. We envisioned that these requirements could be met by implementing the concept of VRT as a Virtual Reality (VR) application and named the device “Salzburg Visual Field Trainer” (SVFT). The advantages over existing VRT systems is that SVFT is compact, portable and comfortable to wear, making the training device ideal for both randomized controlled trials (RCT) and clinical usage during rehabilitation. In combination with the EFA, which provides highly accurate and reliable visual field assessment, another crucial aspect for experiments with the SVFT regarding potentially ameliorating effects of VRT is enabled. That is the unprecedented exact positioning of training stimuli on the patients’ individually shaped border areas. In the following, we describe the device and present a proof-of-concept study of its reliability, which was assessed in a sample of 40 healthy participants.

## Materials and methods

The SVFT was developed using Unity^®^ (Unity Technologies, 2019) with a Google^®^ Cardboard (Google LLC, 2019b) solution, consisting of ZEISS^®^ VR ONE Plus goggles (Carl Zeiss AG, Oberkochen, Germany) in combination with Celly^®^ Bluetooth^®^ remote controls (Celly S.p.A., Vimercate, Italy) and Samsung Galaxy S9^®^ smartphones (Samsung, Seoul, South Korea), running on the Android^®^ Pie operating system (Google LLC, 2019a) (see Fig. 2).

**Figure 2.**
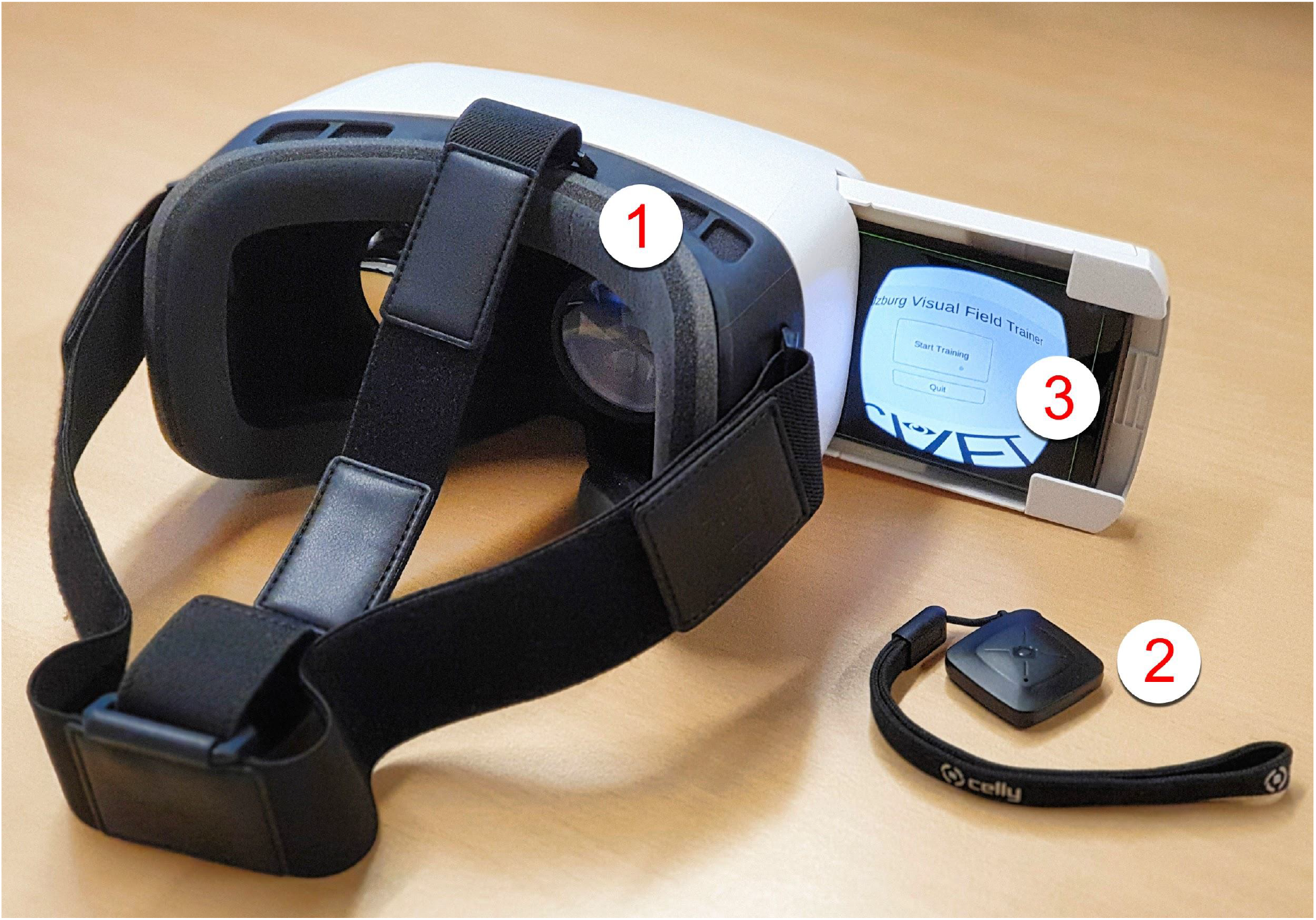
Setup of the the “Salzburg Visual Field Trainer” (SVFT) illustrating Google^®^ Cardboard based solution (1) ZEISS^®^ VR ONE Plus, (2) Celly^®^ Bluetooth^®^ remote control and (3) Samsung Galaxy S9^®^ smartphone (SM-G960F/DS). The smartphone runs software coded in Unity^®^ on Android^®^ Pie operating system. By clicking on the Bluetooth^®^ remote control during training, the user gives feedback to the system, which enables main menu navigation and adapts the position of the presented stimulus during the course of training. Thus, the presented stimuli keep being displayed in the individual transition zone of the user’s intact and defect visual field after potential improvements stemming from neuronal reconnection.

ZEISS^®^ VR ONE Plus houses two aspherical biconvex lenses with +32.5 diopter, enabling the patient to accomodate on the smartphone display, which is inserted in a front tray of the head mounted system in a distance of 44 mm.

### Functionality of the SVFT

The entire input required during the training process can be controlled from the Bluetooth^®^ remote control (consisting of one pressable button), allowing the smartphone to be permanently fixed to the ZEISS^®^ VR ONE Plus socket. Through a side opening, the smartphone can also be charged while it is inserted in the virtual reality goggles. With a double click on the remote control, the SVFT application can be started directly from the smartphone’s standby mode. Then, one single click is required to start training. Subsequently, single clicks during training act as patients’ behavioral feedback, which is required for the stimulus adaptation procedure (see below for details) during rehabilitation sessions. When training is complete, SVFT closes automatically, allowing the smartphone to fall back autonomously into standby mode.

Similar to conventional VRT software on Personal Computers, the patient fixates her/his gaze on a fixation cross in the middle of the screen. Bright light stimuli with approx. 1,000 cd/m^2^ and a size of 3° appear for 750 ms in randomized order on a dark background (see Fig. 3). The presentation of each stimulus is interrupted for 2000 ms (plus a randomized duration of up to 1500 ms) to give the patient time to react to the displayed stimulus via remote control. Training time duration and all other settings mentioned above can be adapted by qualified personnel, allowing a flexible optimization of the therapy setting.

**Figure 3.**
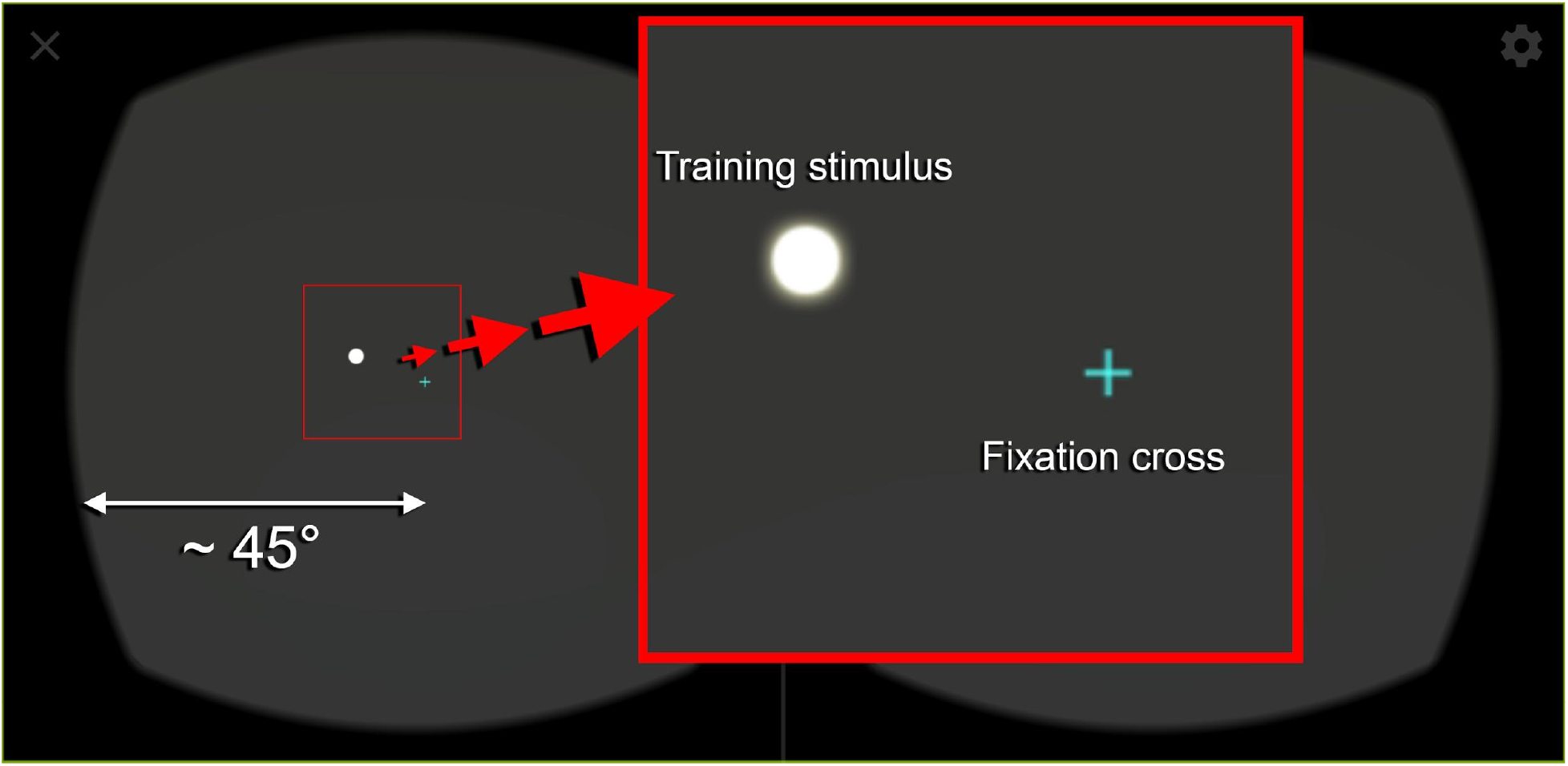
Exemplary stimulus presentation displayed during training with the “Salzburg Visual Field Trainer” (SVFT). Single bright stimuli are presented on a dark background with a centrally placed fixation cross. The digitally enlarged section (red square) shows the fixation cross - which the patients must fixate continuously during therapy - and a training stimulus on an exemplary position in more detail. Based on system settings - which are configured by a qualified person and assessed e.g. with the help of the EFA beforehand - stimuli are displayed randomly along the user’s individual transition zones between intact and defect visual fields. The ZEISS^®^ VR ONE Plus offers a stereoscopic field of view of approx. 90°.

Patients’ behavioral feedback is necessary as the theoretical foundation of VRT states that stimulus presentation should lead to reconnection of lesioned cortical areas, resulting in gradual amelioration of damaged visual field areas (e.g. Sabel & Kasten 2000). Therefore, the SVFT runs a stimulus-location adaptation process during rehabilitation, based on user feedback by clicking (or not clicking) the Bluetooth^®^ remote control during the break after every displayed stimulus. If the user detects the stimulus peripherally, its location is moved towards a custom reference point within the central area of the individual visual field defect. This allows the VRT system to automatically adapt to an improving visual field or small inaccuracies from minor gaze deviations from the fixation cross. On the contrary, if the stimulus was not detected, it is moved away from the reference point towards the center of the patient’s intact visual field, balancing out the therapeutic process in case of inaccuracies, misclicks or loss of already gained training progress. Thus, this behavioral procedure produces a steady approximation of the “real” border across the intact and defect visual field and successively compensates for minor gaze inaccuracies of the patients during the training session procedure. An additional potential benefit of this behavioral procedure is that by pressing a button when concentrating on the peripherally displayed stimuli, attention during training may be further improved as the rehabilitation process becomes an interactive task.

Individual stimuli adaptation progress is stored by the SVFT and retrieved when the next training session starts. Optionally, progress is documented but changed stimuli coordinates are deleted after training and the initial coordinates positions are used for the next training session. These training settings - like stimuli display location and duration, reference point location, break duration between stimuli and total training duration - are fully customizable. Additionally, based on individual diagnosis, parameters like stimulus or fixation cross size and brightness are adjustable to the needs of the patient.

To allow researchers and healthcare specialists tracking the training progress of patients, SVFT periodically saves the current training progress into a protocol file. This is done multiple times - which is customizable - during (in case the training is cancelled) and at the end of each training session. For each training session, a new file is created including time and date, overall session runtime and the number of feedback clicks on the remote control. These parameters allow researchers and healthcare professionals to track technical problems, as well as user compliance and general behavior during the therapy sessions. Additionally, the current locations of the training stimuli are saved, facilitating comparisons with previous sets of stimuli locations and therefore investigations into the patients’ therapy progress are enabled.

### Technical evaluation of the SVFT

The crucial point of a VRT based training device is a reliable and precise presentation of the training stimuli across the individually shaped border area in the patients’ visual field. This central aspect depends on two factors. First, the patient is required to fixate continuously on the central fixation cross in order to perceive the presented stimuli correctly from a parafoveal position accordingly. Because patients expect a therapeutic effect from training with SVFT, they are - based on our clinical experience - usually highly motivated to conduct the training procedure as accurately as possible. Second, the individual characteristics of the patients’ visual field defects must be assessed as precisely as possible and - most importantly - presented stimuli must be placed exactly across the patients’ individual border area during the training procedure. This crucial factor is achieved as follows: In a first step the patients’ visual fields are diagnosed with the help of the EFA. Due to its eye-tracking-based adaptation process, highly accurate data regarding the patients’ perimetric status is assessed (Leitner et al. 2021). Thus, this enables - in a second step - individually custom-tailored positioning of test stimuli in the SVFT, based on every patients’ respective visual field properties and the resulting individual characteristics of the border area between intact and defect visual field.

### Experimental design

To investigate whether training stimuli are displayed correctly in their intended locations in the SVFT, we conducted a proof-of-concept study with a group of 40 healthy participants. Accurate displaying of training stimuli in the individually shaped border area of patients is crucial for the potential therapeutic benefit of VRT based approaches. Therefore, we defined the blind spot - a naturally occuring scotoma in the human visual field - as an objective marker for the degree of precision that SVFT displays stimuli in the patients’ visual fields. This experimental approach is based on the blind spot fixation control mechanism in clinically established automated perimeters (such as the Humphrey Field Analyzer from ZEISS). In these devices the retinal natural blind spot serves as an indicator for patients’ reliability to maintain central fixation during visual field diagnosis. This control mechanism is based on the assumption that if a patient reacts too often to stimuli presented in his/her blind spot, the perimetric assessment needs to be repeated in order to ensure validity. Because humans are unable to detect stimuli presented in their blind spots (due to missing retinal receptor cells), patients’ reactions to such displayed stimuli suggest low compliance rate, stemming from unreliable central fixation and thus leading to biased visual field test results. The blind spot is classified as an absolute scotoma and is located between approximately 12° medial, 17° lateral and 1° below the horizon in a temporal location from the fovea centralis (Burk & Burk 2011). Using the blind spot as a marker of visual field measurement has a tradition in visual field research (Treisman & Fearnley 1976) and the German Ophthalmological Society (DOG) states that the blind spot should serve in every perimetric examination as a referential scotoma (DOG 2019). We also already used this experimental paradigm successfully to investigate and show the high accuracy of the EFA (Leitner et al. 2021).

We evaluated stimulus presentation monocularly in the SVFT by displaying 15 stimuli in the participants’ individual blind spot locations (“blind spot stimuli”) as diagnosed with the help of the EFA and placed another 85 stimuli in the rest of the normal-sighted visual field (“detectable stimuli”). The 40 healthy participants had a mean age of 23 years (SD = 3) and suffered from no reported eye diseases or other clinically ophthalmologic or neurologic issues. Participants were excluded from the study if dioptre was greater than +3 dpt or if other clinical conditions were reported. Our hypothesis was that if the SVFT displays stimuli as intended - based on visual field test results from our validated perimetric methodology EFA - participants should be able to see (and react accordingly correctly) only to detectable stimuli, but not to blind spot stimuli, which they naturally can not see due to their presentation in their absolute scotomas (see Fig. 4).

**Figure 4.**
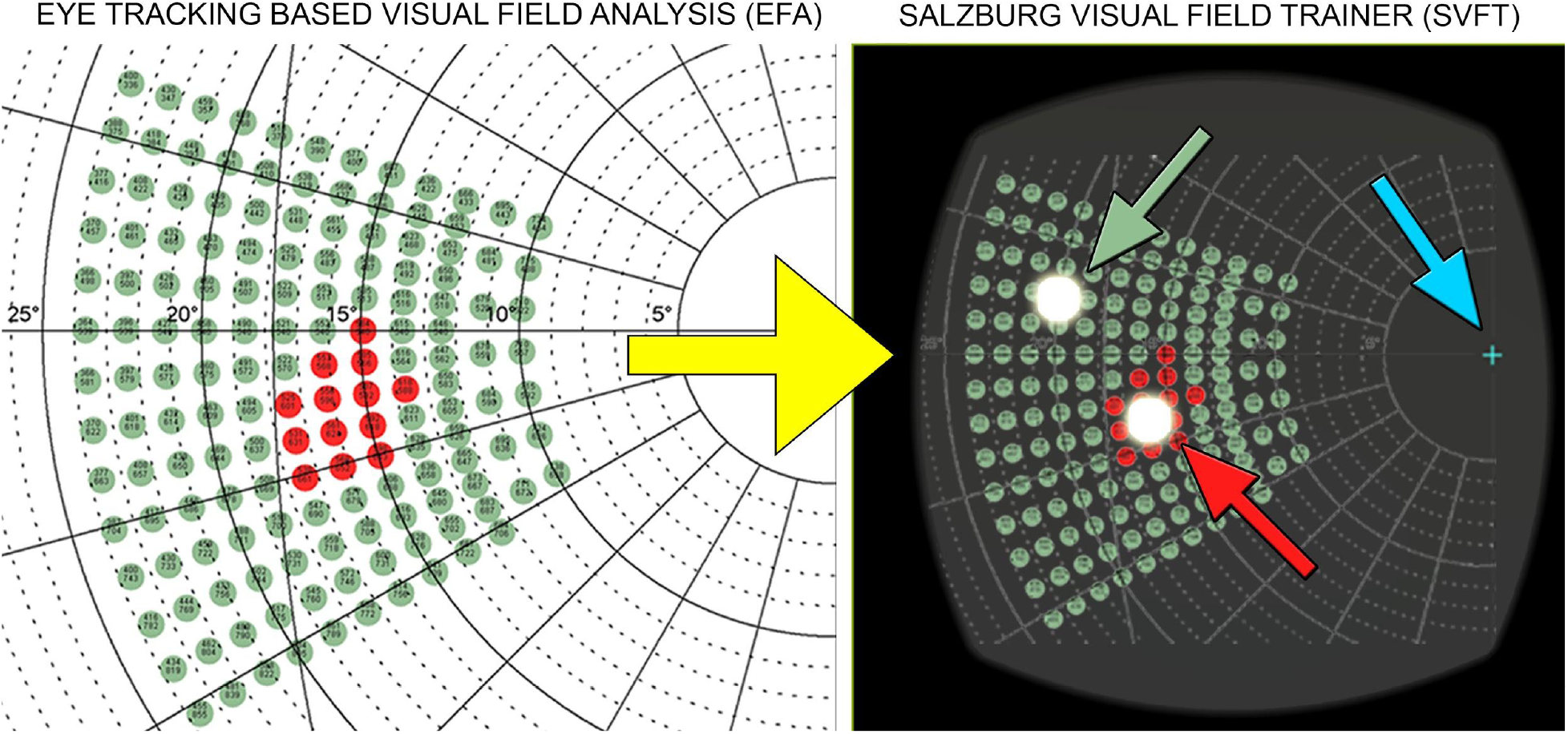
Two-steps experimental design of the study. Left panel (step one): With the help of the validated perimetric methodology “Eye Tracking Based Visual Field Analysis” (EFA) the exact individual location and extent of the blind spots (red cluster) of 40 healthy, normal-sighted participants was assessed. Right panel (step two): Following, the coordinates of the individual blind spots location and extent were transformed (illustrated by the yellow arrow) into the coordinates system of the “Salzburg Visual Field Trainer” (SVFT). Participants were instructed to keep a steady fixation on the central cross (blue arrow) during testing and press the bluetooth button whenever they saw a stimulus in their peripheral vision. From 100 stimuli that were presented, 15 stimuli were displayed in the individually assessed location of the participants’ blind spots (exemplary “blind spot stimulus”: red arrow) and 85 stimuli were displayed around the participants’ blind spots (exemplary “detectable stimulus”: green arrow) in their fully functional visual field.

All stimuli had a size of 0.33° and were displayed for 200 ms, following a time period of 2000 ms to react to the stimulus. A break, randomly ranging from 0 to 1000 ms followed subsequently. Then the next stimulus was presented. Fixation cross color changed randomly after a time period of 3000 ms to 7000 ms. We implemented this color change because we learned from previous experiences that color shifts facilitate fixation of the cross during visual field diagnosis considerably. Similarly as in automated static perimetry assessment, we instructed the participants to react to a set of peripherally presented stimuli via Bluetooth^®^ remote control, while looking steadily at the central fixation cross of the SVFT. Based on visual field assessment with the EFA, we configured the location of the 15 “blind spot stimuli” in SVFT individually for every participant, so the stimuli would fit exactly with the previously assessed participants’ blind spot central location respectively. This means that an “optimal” SVFT test run should result in 85 behavioral reactions (hits) to “detectable stimuli” (correct negative), 0 hits to “blind spot stimuli” (false positive), 15 behavioral misses (missed hits) of undetectable “blind spot stimuli” (correct positive) and 0 missed hits to “detectable stimuli” (false negative). After trial completion, individual participants detection results were automatically stored in a data file on the smartphone, allowing calculation of results regarding sensitivity, specificity, positive predictive value, negative predictive value, hit rate, random hit rate and RATZ-Index for the SFVT (Lenhard & Lenhard 2014).

A total number of 150 stimuli were randomly presented to the participants. The first 25 stimuli were used to accustom the participants to the experimental procedure. Therefore, no “blindspot stimuli” were presented in this block and data was excluded from analysis. The last 25 stimuli were used as a buffer for saving results in the SVFT, hence no “blindspot stimuli” were displayed in this phase of the experiment either and data was also excluded from the analysis. Consequently, testing with the SVFT resulted in a mean total trial duration of 6 minutes and 45 seconds.

Because the EFA and the SVFT are based on two different 2-dimensional coordinate systems, visual field data results from the EFA - which is the basis for the correct placement of the stimuli in the patient’s visual field - were converted into SVFT specific dimensional properties. Based on the users’ viewing distance to the display screens in the EFA (400 mm) and the SFVT (44 mm) (1 retinal angle degree being 1.54 angle degree in the SVFT, based on specifications of the ZEISS^®^ VR ONE Plus goggles) every stimulus (s) location (x and y coordinate from the EFA diagnosis) can be converted from the EFA to the SFVT as follows: 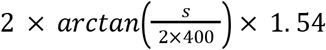

## Results

4000 of 6000 displayed stimuli were used for calculations (see above for details); in total 588 stimuli (mean of the subject = 14.7; SD = 0.6) were recorded as true (correct) positive (TP), 28 stimuli (mean of the subject = 0.7; SD = 1.4) as false positive (FP), 12 stimuli (mean of the subject = 0.3; SD = 0.6) as false negative (FN) and 3372 stimuli (mean of the subject = 84.3; SD = 1.4) as true (correct) negative (TN). This translates to 14.7% correct non-reactions (optimum: 15%), 0.7% false non-reactions (optimum: 0%), 0.3% false reactions (optimum: 0%) and 84.3% correct reactions (optimum: 85%) to displayed stimuli during the presented experiment with the SVFT. The calculation of accuracy (ACC) results in a value of 99.0% 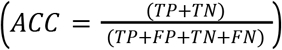

Table 1 shows further results, including sensitivity (SEN), specificity (SPE), positive predictive value (PPV), negative predictive value (NPV), hit rate (HR), random hit rate (RHR) and RATZ-Index (RATZ). SEN denotes the ratio of stimuli correctly identified as “blind spot stimuli”. SPE is the ratio of stimuli correctly identified as “detectable stimuli”. PPW is the probability that a “blind spot stimulus” is correctly identified and NPW is the probability that a “detectable stimulus” is correctly identified. HR denotes the ratio of correct classifications in “blind spot stimuli” and “detectable stimuli”. RHR describes the probability to respond to “blind spot stimuli” by chance. RATZ denotes the increase of HR compared to RHR (Lenhard & Lenhard 2014).

**Table 1.** Test results

| Table 1. Test results |  |  |  |  |  |  |
| --- | --- | --- | --- | --- | --- | --- |
| *SEN* | *SPE* | *PPV* | *NPV* | *HR* | *RHR* | *RATZ* |
| .980 | .992 | .955 | .996 | .990 | .742 | .976 |

*Notes. SEN: sensitivity; SPE: specificity; PPV: positive predictive value; NPV: negative predictive value; HR: hit rate; RHR: random hit rate; RATZ: RATZ-Index.*

## Discussion

In the present study we validated our newly developed virtual reality based rehabilitation methodology “Salzburg Visual Field Trainer” (SVFT), which is based on the principles of “Visual Restitution Therapies” (VRT). Results of this proof-of-concept study indicate that the SVFT and our developed methodology of translating perimetric test results from the “Eye Tracking based Visual Field Analysis” (EFA) to the SVFT is highly reliable and accurate, resulting in precise display of visual stimuli in predefined areas of the users’ visual field. This process is crucial for a potential therapeutic benefit of VRT based instruments, as such methodologies and rehabilitation procedures must be designed to display light impulses as accurately as possible across desired retinal areas (and hence stimulate the appropriate “residual” cortical areas) in the patients’ visual fields. Thus, our data indicates that we created a feasible and exact virtual reality training device and procedure that can help both to facilitate therapy for patients and contribute to the ongoing scientific discussion regarding the “true” effectiveness of VRT. Furthermore, the results indicate that the participants adhered to the experimental instructions and looked principally at the central fixation cross during testing procedure. Because other neuropsychological approaches (such as compensatory based rehabilitation) also depend on exact and precise illustration of training objects and stimuli in the patients’ visual fields, our results indicate that the technical setup of the SVFT can be used for these methodologies as well.

Compared to traditional personal computer and monitor based VRT approaches, our virtual reality based SVFT device offers a number of advantages, which all potentially increase patients’ motivation and compliance to engage in regularly repeated neuropsychological rehabilitation. (1) The training situation becomes significantly more comfortable and convenient as no chin and head rest is necessary. The patient can sit and move freely and perform training e.g., sitting on the couch while listening to music. (2) The general technical and neuropsychological conditions for the training process itself are further improved. Because the distance between the eyes and the presented stimuli is always exactly the same, head movements or imprecise configurations and adjustments conducted by the patient regarding chin and head rest or PC monitor do not affect the crucial aspect of exact stimulus presentation (across the visual border area). (3) The immersive design of the virtual reality goggles makes it possible to rule out external factors like light conditions or other visual distractions in the surrounding environment, potentially confounding the therapeutic effect. After the experiment, the participants of the study confirmed to us that the virtual reality goggles are indeed very comfortable to wear and intuitive to use. We also received similar feedback from the first patients who trained with the instrument and who exhibited very good compliance (for which the ease of application and the comfortability of the instrument is certainly conducive) for the entire duration of the training (up to 6 month).

Obviously, a central aspect in VRT-based methods and other neuropsychological approaches is user compliance. Patients need to steadily fixate the centrally presented cross in order to enable visual stimulation of respective cortical areas from peripherally displayed stimuli. Because in the current version of the SVFT there is no technical possibility to control for central fixation during the experiment, we emphasized the significant importance of maintaining a steady fixation to the participants. We were aware that there are some behavioral approaches to “control” for central fixation, such as naming out loud changes of the color of the fixation cross. However, we decided against this concept in our proof-of-concept study because we did not want to distract participants unnecessarily during the experiment. Furthermore, based on our clinical experience, we can state that these control methodologies do not prevent patients reliably (if they really plan to do so) from performing saccades in order to detect more stimuli. Instead, we concentrated strongly on participants’ compliance and in order to do so we chose only undergraduate psychology students, who we educated extensively and most accurately about the experimental design and the importance to comply with the experimental instructions. Results from the proof-of-concept study indeed indicate that the participants’ compliance was high and they acted during testing as instructed. Regarding future patients - who will train with the SVFT to potentially improve their defect visual field - the issue of compliance will tend to be a smaller problem as in the experimental condition with healthy participants. The reason is simply because patients have a strong natural interest in achieving significant positive therapeutic effects. Compensatory actions, such as the performance of saccades, would run counter their motivation to improve their visual field functionality.

We also learned from the present proof-of-concept study that we will focus - especially in the beginning of the upcoming clinical trials with the SVFT - not only on patients’ compliance but also on patients’ motivation. This is because patients - in order to achieve a potentially measurable positive therapeutic effect - need to train self-reliantly with the SVFT for 6 months, 6 days a week, twice a day for 30 minutes, which requires a high level of motivation. Therefore, it is crucial to help these patients to keep their training motivation as high as possible. We believe that this can be achieved with regular feedback and contact. Although we know from clinical practice that neurological patients are usually highly motivated to improve their deficits, it is essential to pay attention to the patient’s motivation to carry out the therapy as good as possible. From participants’ feedback we additionally learned that the VR goggles are indeed comfortable to wear and that the functionality of SVFT is easy to grasp. This further indicates that - with the help of SVFT - patients will be enabled to conduct independent rehabilitation in a comfortable and uncomplicated way over a long period of time, which will ultimately be essential in order to state robust evidence for any potential effects of VRT or other neuropsychological interventions.

### Conclusion

Our findings indicate that the SVFT can help - in combination with our recently developed and validated perimetric methodology EFA (or any other highly precise and reliable visual field assessment methodology or instrument) - to investigate the true effects of VRT based approaches (or other approaches) and the underlying mechanisms of training-related neuroplasticity in early regions of the visual cortex.

## Data Availability

Data is available from the corresponding author upon reasonable request

## Acknowledgement

The authors would like to thank Christian Kebsak for ophthalmic advice and counseling and Luise Zellner & Leoni Bernstorf for participants management, data acquisition, data analysis and support during the experiment.

## Contribution

Conception and Design: Leitner, Hawelka / Data Acquisition: Leitner, Gütlin / Analysis and Interpretation of Data: Leitner, Hawelka, Gütlin / Writing Publication: Leitner, Hawelka, Gütlin / Critical Revision of Publication: Leitner, Hawelka, Gütlin / Supervision: Hawelka / Resources: Hawelka / Technical Administration and Support: Leitner, Gütlin

## Patient involvement

Patients were not directly involved in the design of this study. Written informed consent was obtained from all participants who received ECTS credits or 10€ for participation in the study. The study design was approved by the ethics committee of the University of Salzburg.

## Data availability

Data is available from the corresponding author upon reasonable request

## Financial support

FWF Austrian Science Fund, Vienna, Grant Number: P31299

## Competing interests

The sponsor or funding organization had no role in the design or conduct of this research No conflicting relationship exists for any author

## Notes

All authors declare that they have no conflict of interest and approve the manuscript

### Competing Interest Statement

The authors have declared no competing interest.

### Author Declarations

The following study was reviewed and approved by the ethics commission of the University of Salzburg (Reference No. 39/2018)

